# Short-term surrogate biomarkers of chronic lesion expansion

**DOI:** 10.1101/2023.04.10.23288349

**Authors:** Samuel Klistorner, Michael H Barnett, John Parratt, Con Yiannikas, Alexander Klistorner

## Abstract

**Objectives:** Slow expansion of multiple sclerosis (MS) lesions has been shown to significantly contribute to disease progression. However, accurate assessment of this metric remains challenging. We investigated whether the long-term damage caused by slow-burning inflammation at the rim of chronic MS lesions can be predicted within timeframe of a typical clinical trial, using surrogate imaging markers.

**Methods:** Pre- and post-gadolinium 3D-T1, 3D FLAIR and diffusion tensor images were acquired from 42 patients with MS. Lesion expansion was analysed annually between baseline and 48 months. The volume of chronic lesion expansion was stratified by the degree of tissue damage within the expanding component of the lesion, measured by a progressive volume/severity index (PVSI). Central brain atrophy (CBA) and the degree of tissue loss inside chronic lesions (measured by the change of T1 intensity and MD) were used as surrogate markers.

**Results:** CBA measured after 2 years of follow-up predicted PVSI at 4 years with a high degree of accuracy (r=0.90, p<0.001, ROC area under the curve 0.92, sensitivity of 94%, specificity of 85%). Increased MD within chronic lesions measured over 2 years was also strongly associated with future PVSI (r=0.80, p<0.001, ROC area under the curve 0.87, sensitivity of 81% and specificity of 81%). In contrast, change in lesion T1 hypointensity poorly predicted future PVSI (best sensitivity and specificity 60% and 59% respectively).

**Interpretation:** CBA and, to a lesser degree, the change in MD within chronic MS lesions, measured over 2 years are reliable and sensitive predictors of the extent and severity of long-term lesion expansion.

## Introduction

Multiple sclerosis (MS) is a chronic inflammatory, neurodegenerative disease of the central nervous system (CNS). Axonal loss is widely regarded as the major cause of irreversible neurological disability in MS. While acute white matter lesions are a principal cause of axonal transection and subsequent axonal degeneration in MS, slow-burning inflammatory demyelination at the edge of chronic lesions (and its Magnetic Resonance Imaging (MRI) equivalent, the expansion of chronic MS lesions) has been implicated as another major factor that contributes to disease evolution, including neurodegeneration, brain atrophy and disability worsening in both the relapsing and progressive stages of the disease.[1][2][3][4][5][6][7] As such, expanding lesions (EL) are a potentially attractive imaging outcome for future clinical trials aimed at neuroprotection.[8]

However, accurate measurement of chronic lesion expansion is challenging, particularly over epochs of two to three years, the typical duration of a treatment trial. Therefore, in the current study, we investigated whether the extent of long-term damage caused by slow-burning inflammation at the rim of chronic MS lesions can be accurately predicted by imaging data routinely collected within timeframe of a typical clinical trial. We examined the performance of several potential imaging surrogates that fulfilled the following criteria: measurement must be fully or largely automated, they must be derived using standard clinical acquisition protocols and they must have previously shown strong association with lesion expansion.

It is understood that lesion expansion (and associated axonal transection) results in retrograde and anterograde (Wallerian) axonal degeneration and eventual loss. Supporting this notion, we recently demonstrated that low-grade inflammation at the lesion rim is associated with considerable (and measurable) damage to, and subsequent degeneration of, traversing axons, leading to loss of periventricular normal appearing white matter (as measured by ventricular enlargement) [9]. In addition, loss of the intra-lesional component of axons that had been transected at the lesion border was shown to cause progressive rarefication within the lesion core, reflecting enlargement of the extracellular space [3][10][11][2].

Therefore, central brain atrophy (CBA), as measured by increased volume of the lateral ventricles, together with isotropic (mean) diffusivity (MD) increase and progressive worsening of T1 hypointensity inside chronic lesions during 1 or 2 years of follow-up, were used in this study as a potential predictive factors of long-term (4 years) lesion expansion[12] [10].

## Method

The study was approved by University of Sydney and Macquarie University Human Research Ethics Committees and followed the tenets of the Declaration of Helsinki. Written informed consent was obtained from all participants.

### Subjects

Patients diagnosed with RRMS according to the revised McDonald 2017 criteria[13] who were enrolled in an existing longitudinal study of MS-related axonal loss and who completed 5 years follow-up were included in the study. Due to significant acceleration of brain atrophy, which becomes exponential in the sixth decade of life[14] [15], only patients younger than 55 years old were included in the study.

Patients underwent MRI scans and clinical assessment at 0, 12, 24, 36 and 60 months. The main analysis was performed between 12 months (termed “baseline”) and 24, 36 or 60 months (termed “1, 2 and 4 years follow-up”), while scans performed at 0 months (termed “pre-study scans”) were used to identify (and exclude) newly developed lesions in the first 12 months of the study.[3]

### MRI protocol and analysis

MRI scans were acquired on a 3T GE Discovery MR750 scanner (GE Medical Systems, Milwaukee, WI) with the following sequences:: pre- and post-contrast (gadolinium) sagittal 3D T1, FLAIR CUBE and diffusion weighted MRI. Specific acquisition parameters and MRI image processing methods are presented in the Supplementary material.

JIM 9 software (Xinapse Systems, Essex, UK) was used to segment individual lesions at baseline and 4 years of follow-up (using co-registered FLAIR CUBE images). Lesion activity during those 48 months was analysed using custom-built software that identifies both the stable and expanding components of chronic lesions (adjusted to correct for brain atrophy-related displacement of lesions at follow-up [3]), and new lesions, as described previously.[16]

The level of axonal damage in the expanding component of ELs is typically moderate, though significant variation exists[3]. We therefore stratified the volume of chronic lesion expansion by the degree of associated tissue damage by introducing a progressive volume/severity index (PVSI), computed by multiplying the volume of expansion by the change of MD in corresponding voxels, as described previously.[9] PVSI between baseline and 4 years was used in this study as a normalised measure of chronic lesion expansion.

Volumetric change of the lateral ventricles at 1 and 2 years of follow-up was used as a measure of CBA.[17][18] [9] It was calculated by multiplying baseline ventricle volume (as segmented by SIENAX) by the percentage ventricular volume change (as calculated by VIENA, an FSL tool).[19]

The degree of lesional tissue damage was determined by measuring the increase of Mean Diffusivity (MD)[20] or T1 hypointensity inside the lesion core between baseline and 1 or 2 years of follow-up [10] [21]. This, however, should not be confused with MD change in the expanding part of the lesion (as used to determine PVSI, described above).

### Statistical analysis

Statistical analysis was performed using SPSS 22.0 (SPSS, Chicago, IL, USA). Pearson correlation coefficient was used to measure statistical dependence between two numerical variables. For partial correlation, data was adjusted for age, gender, disease duration, baseline lesion volume and ventricular size. P < 0.05 was considered statistically significant. Shapiro–Wilk test was used to test for normal distribution. To investigate how well each factor predicts lesion expansion after 4 years of follow-up, we performed a forward stepwise linear regression analysis that included age, gender, disease duration, baseline DMT, baseline lesion volume and CBA (or change of lesional MD or T1 hypointensity). Longitudinal changes were assessed using paired two-sample t-test. Receiver operating characteristic (ROC) curve analysis was performed to determine sensitivity and specificity of CBA and MD/T1 hypointensity change inside lesions at 2 years in predicting future lesion expansion.

## Results

Forty-two consecutive MS patients (24F, 18M, mean age 40.7±8.1, median EDSS 1, range (1-3) and disease duration 5.6+/-4.7 years) with 5 years of follow up satisfied the inclusion criteria and were enrolled in the study. Six patients were maintained on lower efficacy treatment (injectables, such as interferon and glatiramer acetate, teriflunomide and dimethyl-fumarate)[22], while 21 patients were receiving higher efficacy drugs (fingolimod, natalizumab, and alemtuzumab)[22] during the study period. Two patients were treatment-free, while 13 patients changed treatment category between baseline and follow-up visits.

There was a significant (p<0.001) increase of total lesion volume during the 4 year follow-up period (Table 1), 80% of which was due to the expansion of chronic lesions; and 20% to new lesions.

|  | Total lesion volume<br>(mm<sup>3</sup>) | Lesion expansion<br>(mm<sup>3</sup>) | New lesions<br>(mm<sup>3</sup>) |
| --- | --- | --- | --- |
| Baseline | 5556±5013 | | |
| Follow up (4 years) | 6283±5408 | 584±866 | 143±128 |

### 1. Early CBA predicts subsequent lesion expansion

There was significant association between CBA measured after 1 and 2 years of follow-up and PVSI of chronic lesions measured at 4 years. However, the correlation was considerably stronger for 2 years (correlation coefficient r=0.74 and r=0.90 respectively, p<0.001 for both) (Fig.1, upper row).

**Figure 1.**
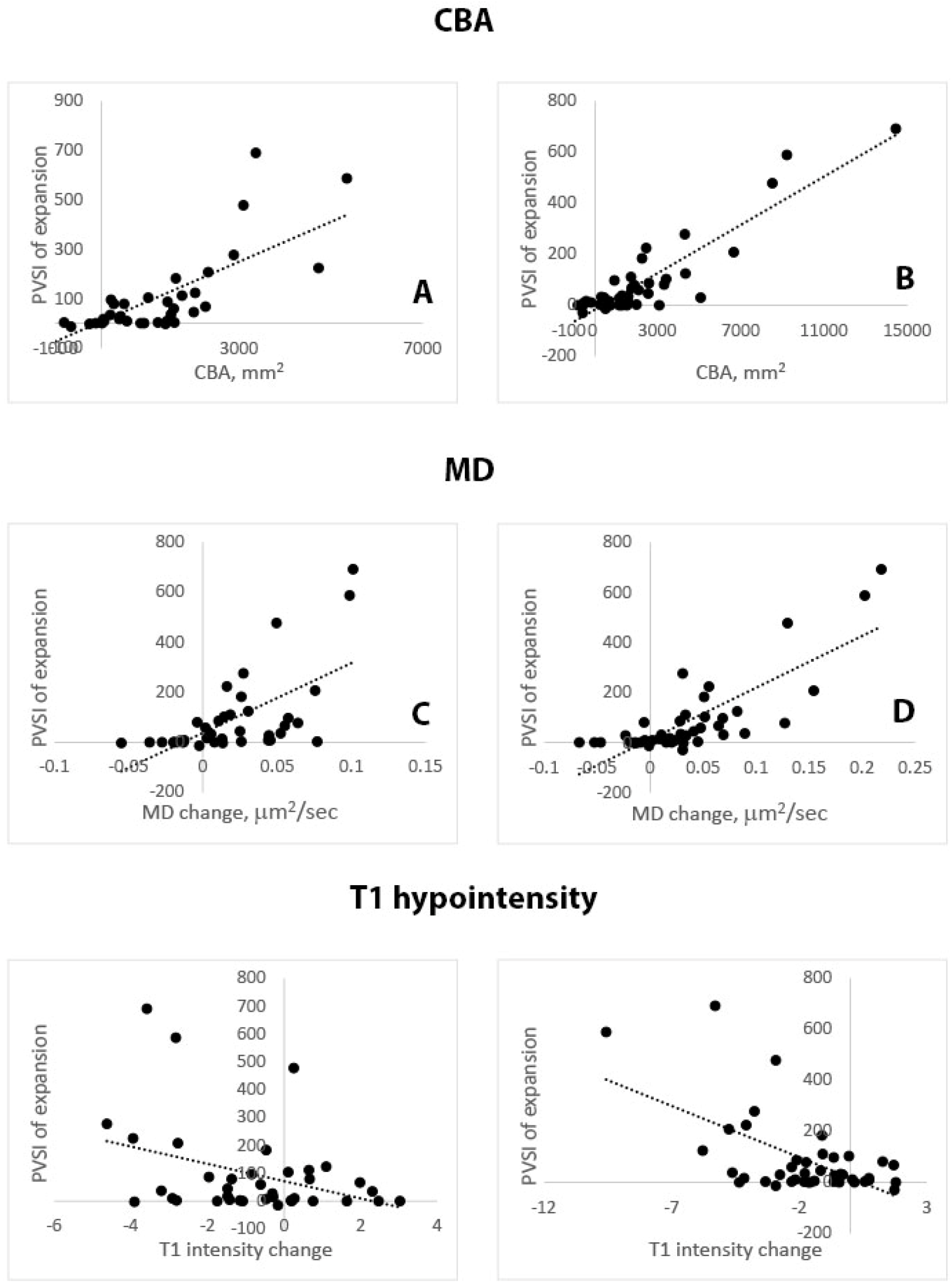
Correlation of PVSI at 4 years of follow-up with CBA, MD and T1 hypointensity at 1 year (left column) and 2 years (right column).

Multiple linear regression analysis, which, in addition to CBA included age, gender, disease duration, baseline DMT and baseline lesion volume only marginally increased the power of the model to predict long-term PVSI change (r=0.79 and 0.92, p<0.001 for both).

Based on the distribution of the PVSI values in our patient cohort, it was assumed that an index above 50 is indicative of significant damage caused by the expansion of chronic lesions. Consequently, based in this cut-off patients were separated into expanding vs non-expanding groups.

Receiver operating characteristic (ROC) analysis of PVSI using 2 years CBA follow-up data as a predictor yielded an area under the curve of 0.92. CBA cut-off of 1670mm^3^ predicted long-term (i.e. 4 years) damage from chronic lesion expansion with a sensitivity of 94% and specificity of 85% (Fig.2a).

**Figure 2.**
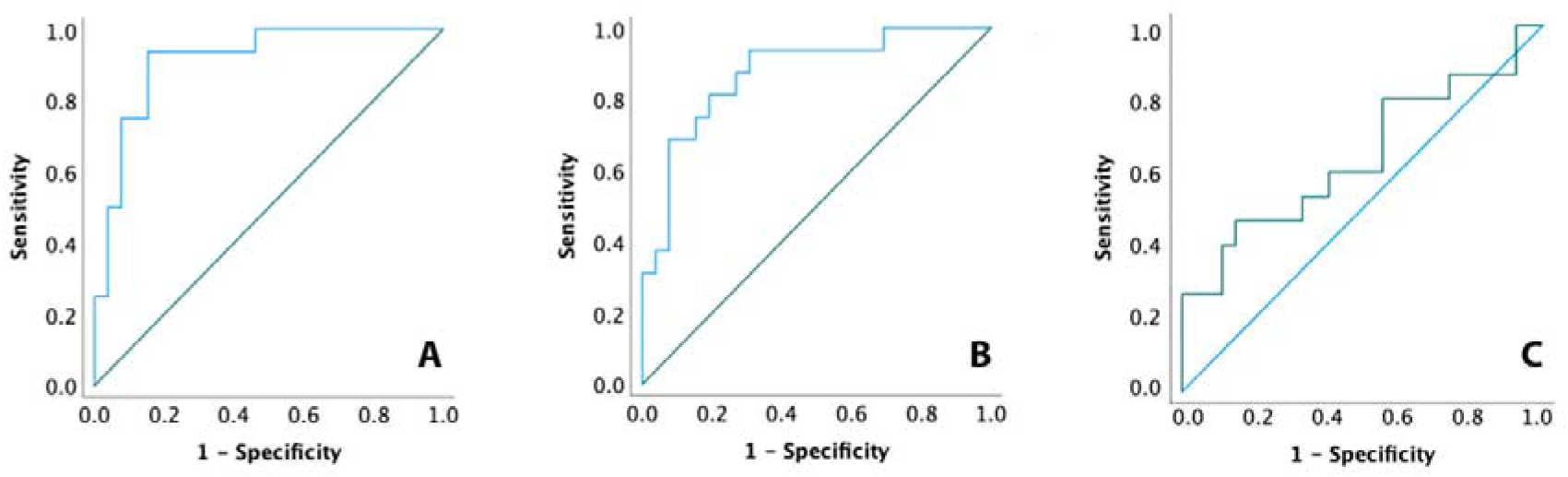
ROC analysis of CBA (A), MD (B) and T1 hypointensity (C).

### 2. Early tissue rarefication in chronic lesions and subsequent lesion expansion

#### a. MD

The PVSI of the expanding part of chronic MS lesions measured over 4 years correlated significantly with increased MD within the core of chronic lesions measured after 1 and 2 years of follow-up. Again, the correlation for 2 years remained much stronger (correlation coefficient r=0.62 and 0.80 respectively, p<0.001) (Fig.1, middle row).

Multiple linear regression analysis of MD change including the baseline factors mentioned above only slightly improved the predictive power of the model (r=0.67 and 0.83 for 1 and 2 years of follow-up respectively, p<0.001 for both).

Patient-based ROC analysis of PVSI using the same criteria to separate patients into groups as described for CBA demonstrated an area under the curve = 0.87. Increased MD > 0.032 μm^2^/sec within chronic lesions during 2 years of follow-up predicted long-term lesion expansion with a sensitivity of 81% and specificity of 81%.

#### b. T1 hypointensity

There was also an association between change of T1 intensity inside chronic lesions at 1 and 2 years of follow-up and PVSI of the expanding part of the chronic lesions measured at 4 years (correlation coefficient r=0.36 and 0.59 respectively, p=0.02 and p<0.001 respectively). While significant, this correlation was moderate at best (Fig.1, bottom row).

Multiple linear regression analysis only marginally increased the predictive power of the model (r=0.40 and 0.61, p<0.001).

ROC analysis of T1 intensity demonstrated poor separation of patients with expanding vs non-expanding lesions (ROC area under the curve = 0.64, with the best sensitivity of 60% and specificity of 59%.)

## Discussion

Slow-burning demyelination at the edge of chronic lesions and associated gradual lesion expansion has been implicated as an important pathological substrate for clinical progression[23],[24]. While progressive loss of axons is recognised as the direct cause of disability worsening[25],[1],[26] in MS, quantitative assessment of brain damage caused by axonal transection at the expanding rim of chronic lesions is not well established.

Several recent studies have attempted to address this issue. The incidence of slowly expanding lesions (SEL), detected based on Jacobean non-linear transformation has been used in a number of investigations [2],[11],[27]. While largely automated, this technique is limited to SEL count and does not allow accurate quantitative assessment of the brain tissue damage caused by the slow-burning inflammation. Manual lesion delineation, employed in other studies, while more quantifiable, requires long periods of follow-up (due to very slow lesion progression and inherent variability of manual masking), which is beyond the duration of a typical clinical trial and may delay the application of potentially beneficial treatments [3],[28].

Therefore, in the current study we tested surrogate markers of lesions expansion that satisfied the following criteria: automated measurement, derived using standard clinical imaging protocols and a previously demonstrated strong association with lesion expansion.

Our results show that CBA, measured over 2 years with widely adopted and fully automated algorithms (SIENAX and VIENA), can predict long-term lesion expansion with a high degree of certainty, explaining >80% of between-patient variability in normalised lesion expansion at 4 years. ROC analysis also demonstrated a high sensitivity and specificity of CBA in separating patients into expanding lesion vs non-expanding lesion groups obtained after two years of follow-up with a sensitivity of 94% and specificity of 85%. One year data, however, was significantly less reliable.

We recently demonstrated a close link between expansion of chronic lesions and CBA in a four-year longitudinal study of patients with RRMS[9]. This study also revealed that the contribution of white matter loss caused by axonal transection at the rim of expanding lesions accounts for more than 75% of CBA variability. The substantial contribution of lesion expansion to central brain atrophy may reflect the recent observation that expansion of chronic lesions is most apparent in proximity to the ventricles [28]. Therefore, it seems plausible that Wallerian and retrograde degeneration of axons transected at the site of lesion expansion would lead to predominant loss of periventricular white matter and consequently to expansion of the ventricles (i.e. CBA). Axons in periventricular tracts are also considerably longer than those in other white matter areas, such as sub-cortical white matter, and therefore, their transection is likely to have larger effect on periventricular white matter atrophy.

We also investigated the potential use of intra-lesional measures to predict long-term lesion expansion. Pathological studies demonstrated that low-grade demyelination at the rim of mixed active/inactive lesions correlates with axonal injury in their core[29],[26]. Therefore, we examined two biomarkers of lesional tissue rarefication, namely increase of MD and reduction of T1 intensity. We have previously demonstrated a strong association between the degree of chronic lesion expansion and increased isotropic water diffusion (as measured by MD) in the lesion core. [3] Elevated MD inside MS lesions is believed to be related to the enlargement of the extracellular space[30],[31],[32],[33],[34] caused by combination of axonal loss and relatively rigid lesional structure, due to fibrillary gliosis that developed during the acute stage of lesion formation[35]). Low-grade inflammatory demyelination and associated axonal injury occurring at the lesion rim leads not only to degeneration of extra-lesional component of transected axons (and, as a result, contributes to CBA), but also to the attrition of their intra-lesional segments, thereby contributing to increased MD within the lesion). [21],[10] (Fig.3)

**Fig. 3.**
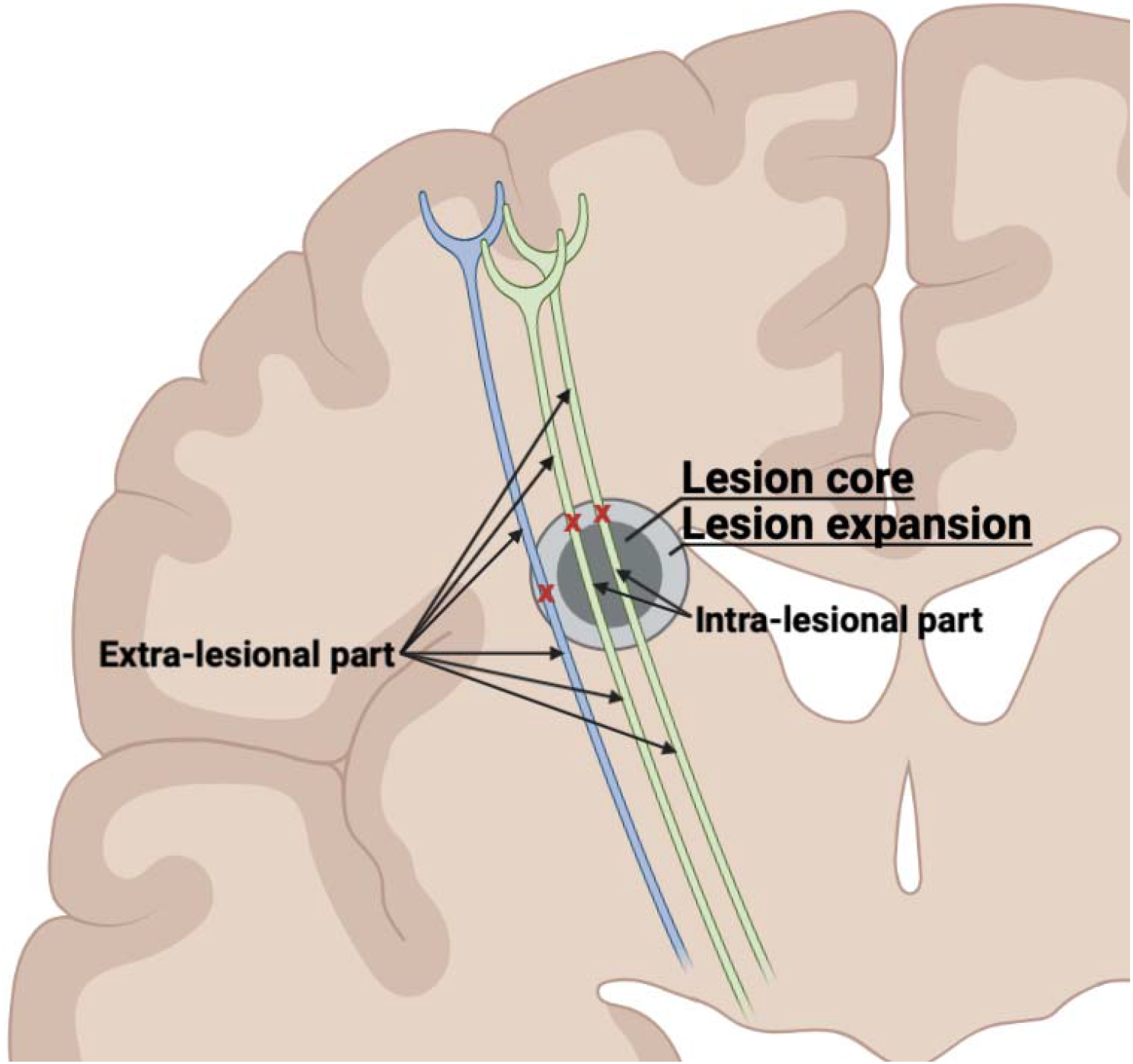
Schematic representation of axonal transection at the rim of a chronic MS lesion. Degeneration of the proximal and distal parts (the ‘extra-lesional’ parts) of all axons transected at the lesion rim (indicated by red crosses) contribute to loss of NAWM and, consequently, to progression of CBA, while only loss of the intra-lesional part of axons that intersect the lesion itself (shown in green) impacts rarefication of tissue inside the lesion (and, accordingly, increased MD/T1 hypointensity).

In addition to MD measures, T1 hypointensity has long been used as an imaging metric that is closely associated with the degree of tissue destruction within MS lesions[36] [37]. Several recent studies have reported that progressive increase of T1 hypointensity (whether by severity or by tissue volume) within pre-existing T2 lesions is linked to lesion expansion. [2] [11] [5][28] Thus, early work by Elliott et al and Calvi et al demonstrated a progressive and significant decrease in T1 intensity within slowly expanding lesions compared to non-expanding lesions[38] [27] and more recent reports confirm that increased (non-gadolinium enhancing) T1 hypointense lesion volume is primarily restricted to chronic (pre-existing) T2 lesions [2][5]

A shared pathomechanism underpinning increased MD and T1 hypointensity within ELs is corroborated by the recent observation that parallel worsening in these metrics follow a periventricular gradient[28].

Based on this body of literature, we speculated that short-term change in these lesional metrics (MD and T1 hypointensity) may provide some guidance as to whether the patient is likely to show tendency for lesion expansion in a longer term.

Our study revealed a relatively strong association between change in MD within the core of chronic lesions at 2 years and degree of long-term lesion expansion. Change of lesional MD explained >60% of variability of chronic lesion expansion between the patients. However, based on ROC analysis, the MD coefficient was outperformed by CBA as a predictor of long-term damage caused by lesion expansion, even after 2 years of follow-up. Furthermore, change of T1 intensity in the lesion core, while significantly associated with severity of lesion expansion, showed only a weak to moderate correlation. As a result, ROC analysis was unable to predict future lesion expansion with a sufficiently high degree of certainty.

The superior performance of CBA (compared to measures of intra-lesional tissue damage) in predicting long-term lesion expansion may relate to the fact that CBA is a global measure that is able to account for all extra-lesional axonal loss that is driven by axons that cross through the lesion core and those that cross exclusively through the zone of lesion expansion. In contrast, change of MD/T1 inside the lesion core is due to degeneration of axons that only pass through the lesion core, representing only a subset of transected axons (Fig. 3).

Limitations of our study include a relatively small sample size that may impact the robustness and generalisability of our conclusions; and preclude assessment of the potential impact of disease modifying therapies on the results of the study. Furthermore, other factors, not considered in this study, such as cortical and spinal cord lesions, may potentially affect some of the measured outcomes, especially CBA.

In conclusion, CBA and, to lesser degree, the change of MD in lesion core, measured within the timeframe of a typical clinical trial, provides a means to quantitatively estimate potential long-term axonal injury caused by slow-burning inflammation at the rim of chronic MS lesions.

## Data Availability

All data produced in the present study are available upon reasonable request to the authors

